# Deep Learning-Based Spatial Profiling Identifies MET-Associated Immune Exclusion and Therapeutic Vulnerability Underlying Racial Disparities in Head and Neck Cancer

**DOI:** 10.64898/2026.10.26.26366494

**Authors:** Xinyu Cui, Fanghui Chen, Ashley McCook-Veal, Yuan Liu, Nicole C. Schmitt, Christine H. Chung, Conor E. Steuer, Dong M. Shin, Nabil F. Saba, Yong Teng

**Affiliations:** Department of Hematology and Medical Oncology, Emory University, Atlanta, GA 30322, USA; Winship Cancer Institute, Emory University, Atlanta, GA 30322, USA; Department of Biostatistics & Bioinformatics, Emory University, Atlanta, GA 30322, USA; Department of Otolaryngology – Head and Neck Surgery, Emory University, Atlanta, GA 30322, USA; Department of Head and Neck-Endocrine Oncology, Moffitt Cancer Center, Tampa, FL 33612, USA; Wallace H. Coulter Department of Biomedical Engineering, Georgia Institute of Technology and Emory University, Atlanta, GA 30322, USA

**Author notes:** **Correspondence:** Yong Teng, PhD, Department of Hematology and Medical Oncology Emory University, Atlanta, GA 30322.

**Keywords:** Head and neck cancer, MET, racial disparities, immune disparities, GigaTIME, Phoenix

## Abstract

Black/African American (BAA) patients with head and neck squamous cell carcinoma (HNSCC) experience more aggressive disease and poorer survival outcomes than White patients; however, the tumor immune microenvironmental features underlying these racial disparities remain poorly characterized. Here, we identify MET as a potential molecular link between immune exclusion and therapeutic response associated with racial disparities in HNSCC. Compared with White-derived tumors, deep learning-based spatial profiling revealed a distinct immune landscape in BAA-derived tumors, characterized by reduced CD8⁺ T cell infiltration, increased immune checkpoint expression, and elevated spatial MET expression. Across independent cohorts, higher MET expression was consistently associated with reduced CD8⁺ T cell infiltration, supporting a relationship between MET-associated tumor biology and an immune-excluded tumor microenvironment. Importantly, in a phase 2 clinical trial cohort treated with pembrolizumab plus cabozantinib, higher predicted spatial MET expression was strongly associated with treatment response and improved clinical outcomes, with the association particularly pronounced among BAA patients. This study establishes a spatially resolved framework linking tumor-intrinsic MET expression with the immune microenvironment and therapeutic response, suggesting that MET-associated biology may contribute to racial disparities in HNSCC progression and treatment outcomes.

## INTRODUCTION

Head and neck squamous cell carcinoma (HNSCC) remains a major global cancer burden, ranking as the seventh most common cancer worldwide with approximately 940,000 new cases and 480,000 deaths in 2022, and long-term survival remains limited despite advances in surgery, radiotherapy, immunotherapy, and targeted systemic treatment^1–3^. In the United States, marked racial disparities in HNSCC outcomes have been recognized for decades, with Black/African American (BAA) patients experiencing higher disease burden and lower survival than White patients^4–10^. These disparities are partly explained by differences in stage at diagnosis, socioeconomic context, healthcare access, and treatment delivery, yet survival gaps can persist after adjustment for neighborhood socioeconomic status and access-related variables^9^. These observations suggest that structural determinants remain central but may not fully explain outcome differences, motivating investigation into tumor-associated molecular and microenvironmental features that differ between BAA and White patients and remain incompletely characterized^10–12^.

Multi-omic studies have begun to address this critical gap by providing a more comprehensive understanding of the molecular and cellular features associated with racial disparities in HNSCC^8,11,13^. Integrative analyses of genomic, transcriptomic, epigenomic, and proteomic data revealed race-associated differences in the frequency of driver gene mutations, including TP53 and FAT1, as well as differences in copy number alterations and DNA methylation patterns between HNSCC tumors from BAA and White patients^12^. Our previous study further identified GSTM1, a xenobiotic-metabolizing enzyme, as a BAA-associated molecular determinant^8^. Elevated GSTM1 expression and copy number gain were associated with poorer survival in BAA patients with HNSCC, highlighting a potential molecular contributor to racial disparities in HNSCC outcomes^8^. Yet these studies rely largely on bulk profiling or single-marker immunohistochemistry, neither of which can resolve how immune and stromal populations, including CD8^+^ T cells and cancer-associated fibroblasts (CAFs), are spatially organized within the tumor^14,15^. Multiplex immunofluorescence (mIF) and spatial transcriptomics can directly characterize these spatial relationships at cellular and molecular resolution^16,17^. However, their substantial tissue, reagent, technical, and instrumentation requirements limit their scalability across large patient cohorts^14,15^. Consequently, how the spatial organization of tumor and immune cells differs between BAA and White patients, and whether these differences contribute to racial disparities in HNSCC, remains poorly understood^11,12^.

To overcome this limitation, we leveraged hematoxylin and eosin (H&E)-stained HNSCC whole-slide images and deep learning-based virtual staining and spatial transcriptomic inference to characterize race-associated differences in tumor microenvironmental organization at scale^18,19^. Applying this approach to The Cancer Genome Atlas (TCGA) HNSCC cohort^20^ and independent cohorts from Emory University, we systematically mapped the spatial distribution and cellular interactions within the tumor microenvironment (TME) and identified distinct immune and molecular features associated with BAA and White patients. Here we report a CD8⁺ T cell-low, checkpoint-high phenotype in BAA-derived tumors, characterized by reduced CD8⁺ T cell infiltration and elevated PD-1/PD-L1 expression compared with White-derived tumors. MET was significantly more enriched in BAA-derived tumors than in White-derived tumors and was spatially associated with reduced CD8⁺ T cell abundance, with this association being more pronounced in BAA-derived tumors. Extending these findings clinically, predicted MET expression, but not predicted PD-L1 expression, was associated with response to cabozantinib (a MET-targeting multikinase inhibitor) plus pembrolizumab (a PD-1 inhibitor) in a phase 2 trial of recurrent or metastatic HNSCC, with the response association appearing more pronounced among BAA patients. These findings implicate MET-associated immune exclusion as a potential mechanism underlying racial disparities in HNSCC and identify MET as a potential therapeutic vulnerability that may be more relevant to BAA patients.

## RESULTS

### HPV-positive and HPV-negative HNSCC exhibit distinct tumor immune microenvironments

To investigate variation in the tumor immune microenvironment of HNSCC, we applied GigaTIME^18^, a deep learning-based virtual staining framework, to H&E whole-slide images from the TCGA HNSCC cohort (**Fig. 1A**). All available formalin-fixed, paraffin-embedded (FFPE) tumor sections with matched clinical metadata were included, yielding 450 unique cases after excluding frozen sections for optimal virtual staining performance. Virtual mIF maps were generated for 20 protein markers^18^, and whole-slide mean marker intensity was quantified within DAPI-thresholded tissue regions. Tumors were stratified as HPV-positive (n = 46) or HPV-negative (n = 404). Quantitative comparison revealed significantly elevated CD8 and cytokeratin (CK) expression in HPV-positive tumors (**Fig. 1B-D**).

**Figure 1.**
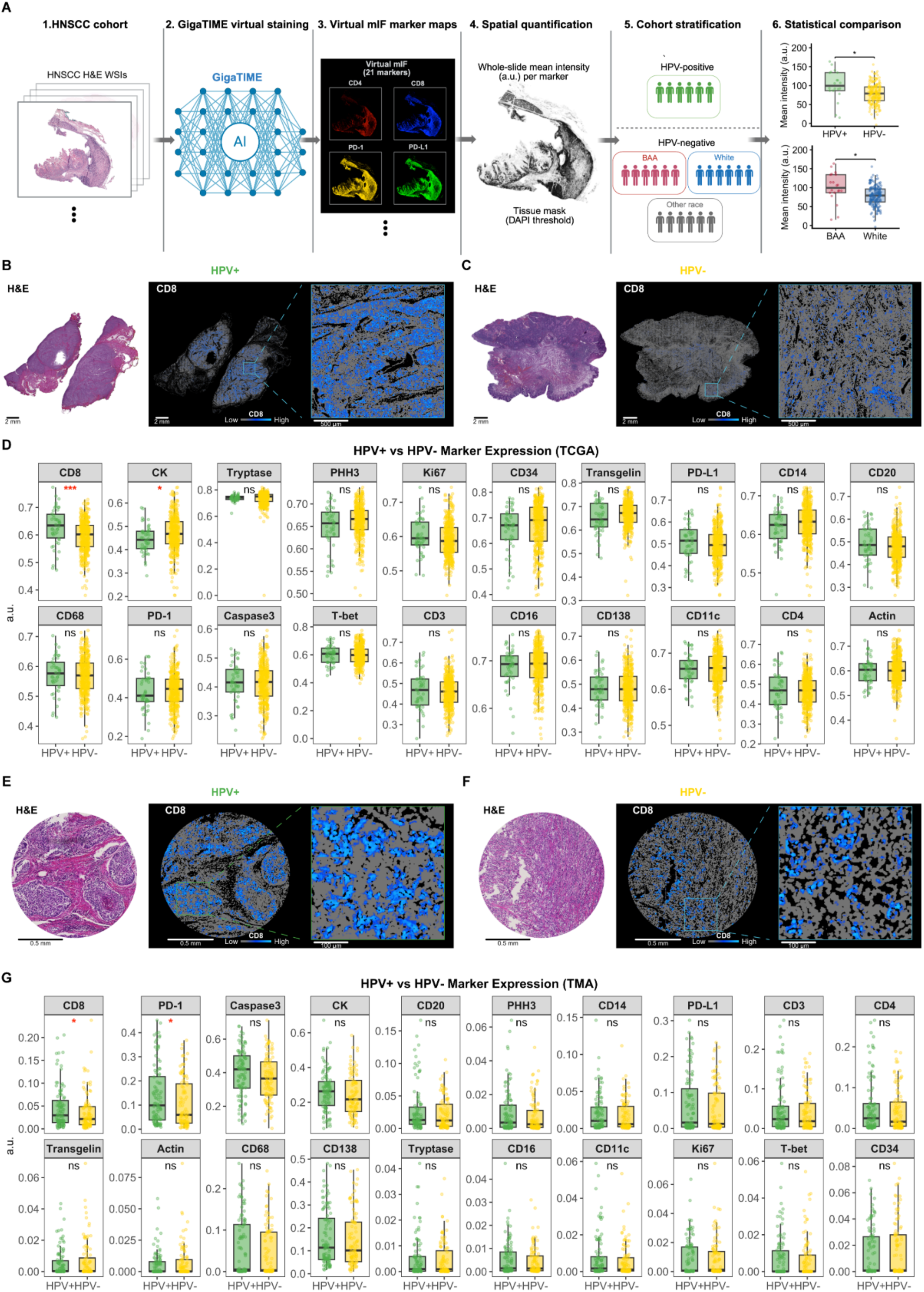
Virtual multiplex immunofluorescence reveals HPV-dependent immune remodeling in HNSCC. (**A**) Overview of the analytical workflow for virtual multiplex immunofluorescence profiling of H&E-stained HNSCC specimens using GigaTIME. (**B, C**) Representative H&E images, whole-slide CD8 virtual staining heatmaps, and magnified CD8-positive regions from an HPV-positive tumor and an HPV-negative tumor in the TCGA cohort. Inset squares indicate 2 mm × 2 mm regions shown at higher magnification. (**D**) Whole-slide mean expression of 20 virtual immunofluorescence markers comparing HPV-positive (n = 46) and HPV-negative (n = 404) HNSCC specimens from the TCGA cohort. (**E, F**) Representative H&E images, whole-slide CD8 virtual staining heatmaps, and magnified CD8-positive regions from an HPV-positive tumor and an HPV-negative tumor in the EMORY TMA cohort. Inset squares indicate 2 mm × 2 mm regions shown at higher magnification. (**G**) Whole-slide mean expression of 20 virtual immunofluorescence markers comparing HPV-positive (n = 100) and HPV-negative (n = 100) HNSCC specimens from the Emory TMA cohort. Scale bars, 2 mm (whole-slide images) and 500 μm (magnified regions). Box plots show median and interquartile range, with individual samples overlaid. ns, not significant, and *p < 0.05, by Wilcoxon rank-sum test.

Next, we constructed an in-house tissue microarray (TMA) from tumor specimens obtained from an Emory University HNSCC patient cohort. The TMA consists of 100 HPV-positive and 100 HPV-negative HNSCC cases with comprehensive clinical and pathological annotations. The same analyses were performed in this independent cohort to validate the TCGA findings. Consistently, HPV-positive TMA cores exhibited significantly higher CD8 expression, with PD-1 also significantly elevated (**Fig. 1E-G**), supporting a more immune “hot” microenvironment in HPV-positive tumors characterized primarily by increased CD8⁺ T cell infiltration.

To integrate these markers into a prognostic framework, we developed a GigaTIME-based risk score using LASSO Cox regression across all 21 markers (**Fig. S1A-B**). The risk score significantly stratified overall survival in HPV-negative patients (log-rank P < 0.0001; **Fig. S1C**) and was significantly higher in BAAs than in White patients (P = 3.7 × 10⁻⁵; **Fig. S1D**). In contrast, the risk score neither stratified survival (P = 0.65; **Fig. S1E**) nor differed between racial groups (P = 0.98; **Fig. S1F**) among HPV-positive patients. Given the limited number of HPV-positive cases available and the absence of race-dependent signal in this subgroup, all subsequent analyses were restricted to HPV-negative HNSCC.

### HPV-negative HNSCC exhibits race-dependent differences in the immune microenvironment

Having established the HPV-negative subgroup as the focus, we next examined whether the tumor immune microenvironment differs between BAA and White patients. After excluding cases from other racial groups (n = 25), HPV-negative tumors were stratified into BAA (n = 41) and White (n = 338) subgroups. Qualitative comparison of representative cases revealed distinct patterns of immune marker expression between the two groups (**Fig. 2A-F**). Whole-slide quantification across 20 markers showed significantly higher PD-1 and PD-L1 expression in BAA-derived tumors, whereas White-derived tumors exhibited significantly higher CD8 expression (**Fig. 2G**). This divergent pattern, characterized by elevated immune checkpoint expression in BAA-derived tumors alongside reduced CD8-associated cytotoxic T cell infiltration, suggests a more checkpoint-enriched and less cytotoxic immune microenvironment in BAA patients.

**Figure 2.**
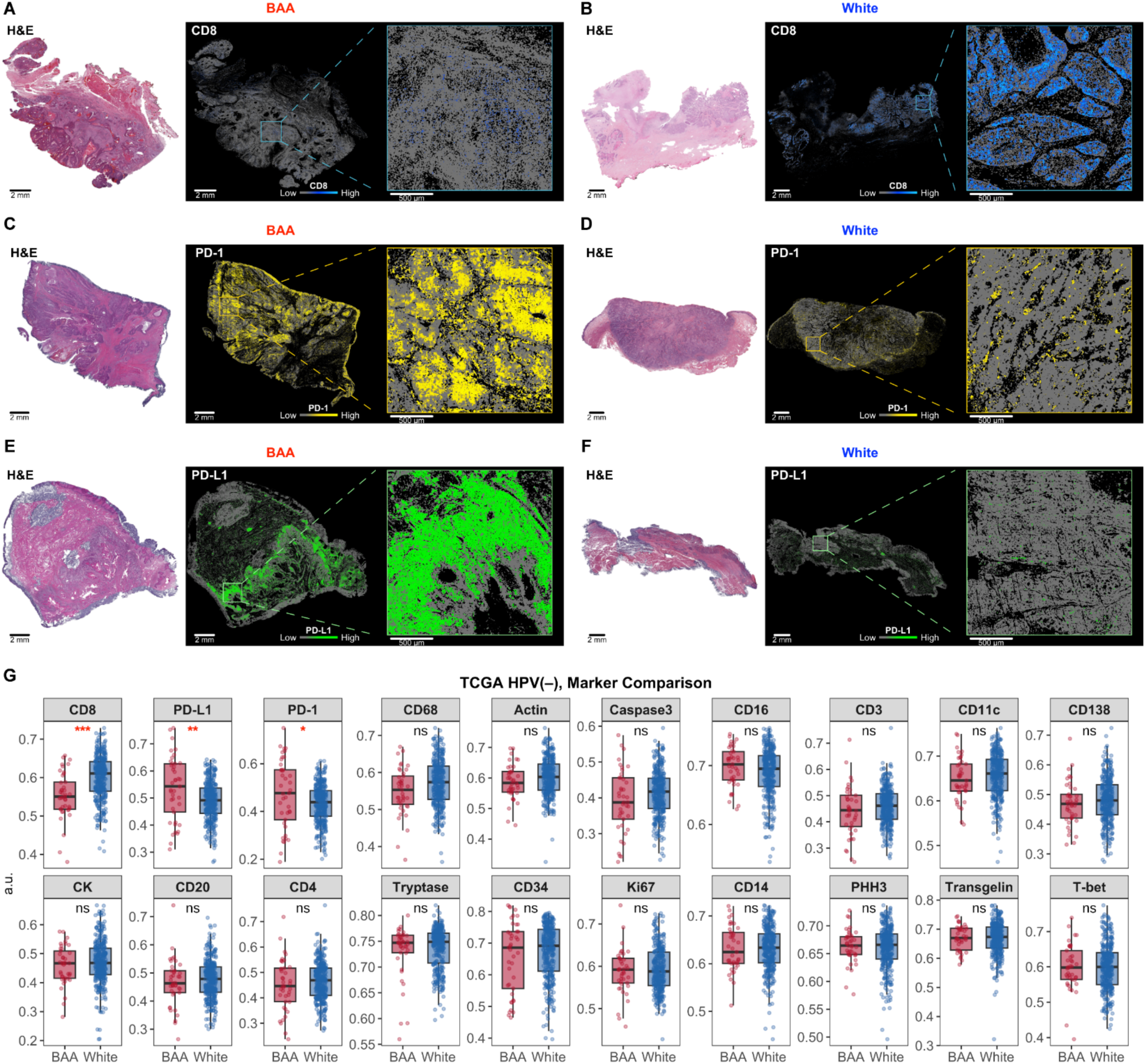
Virtual multiplex immunofluorescence reveals race-dependent immune differences in the TCGA HPV-negative HNSCC cohort. **(A-F)** Representative H&E images, whole-slide virtual staining heatmaps, and magnified regions for CD8 **(A, B)**, PD-1 **(C, D)**, and PD-L1 **(E, F)** in BAA and White HPV-negative tumors. Inset squares indicate regions shown at higher magnification. **(G)** Whole-slide mean expression of 20 virtual immunofluorescence markers comparing BAA and White HPV-negative tumors. Scale bars, 2 mm (whole-slide images) and 500 μm (magnified regions). Box plots show median and interquartile range, with individual samples overlaid. ns, not significant, *p < 0.05, **p < 0.01, and ***p < 0.001, by Wilcoxon rank-sum test.

To validate the race-dependent immune differences identified in the TCGA cohort, we applied the same virtual staining and quantification pipeline to the independently assembled Emory TMA cohort. Representative TMA cores from BAA and White HPV-negative cases showed visually distinct patterns of CD8, PD-1, and PD-L1 expression (**Fig. 3A-F**), consistent with the qualitative differences observed in the TCGA whole-slide images. Quantitative comparison across 20 markers confirmed that BAA-derived tumors again exhibited significantly higher PD-1 and PD-L1 expression, while White-derived tumors showed significantly higher CD8 expression (**Fig. 3G**), reproducing the checkpoint-enriched and less cytotoxic phenotype observed in BAA-derived tumors in the TCGA cohort. To experimentally validate these computationally inferred differences, we performed CD8α IF staining in HNSCC specimens collected through Cancer Tissue and Pathology (CTP) Shared Resource at Winship Cancer Institute of Emory University. (BAA, n = 13; White, n = 13). BAA-derived tumors exhibited significantly lower number of CD8-positive cells than White-derived tumors (**Fig. 3H-J**), confirming race-associated differences in the immune microenvironment of HPV-negative HNSCC across both virtual and experimental staining platforms.

**Figure 3.**
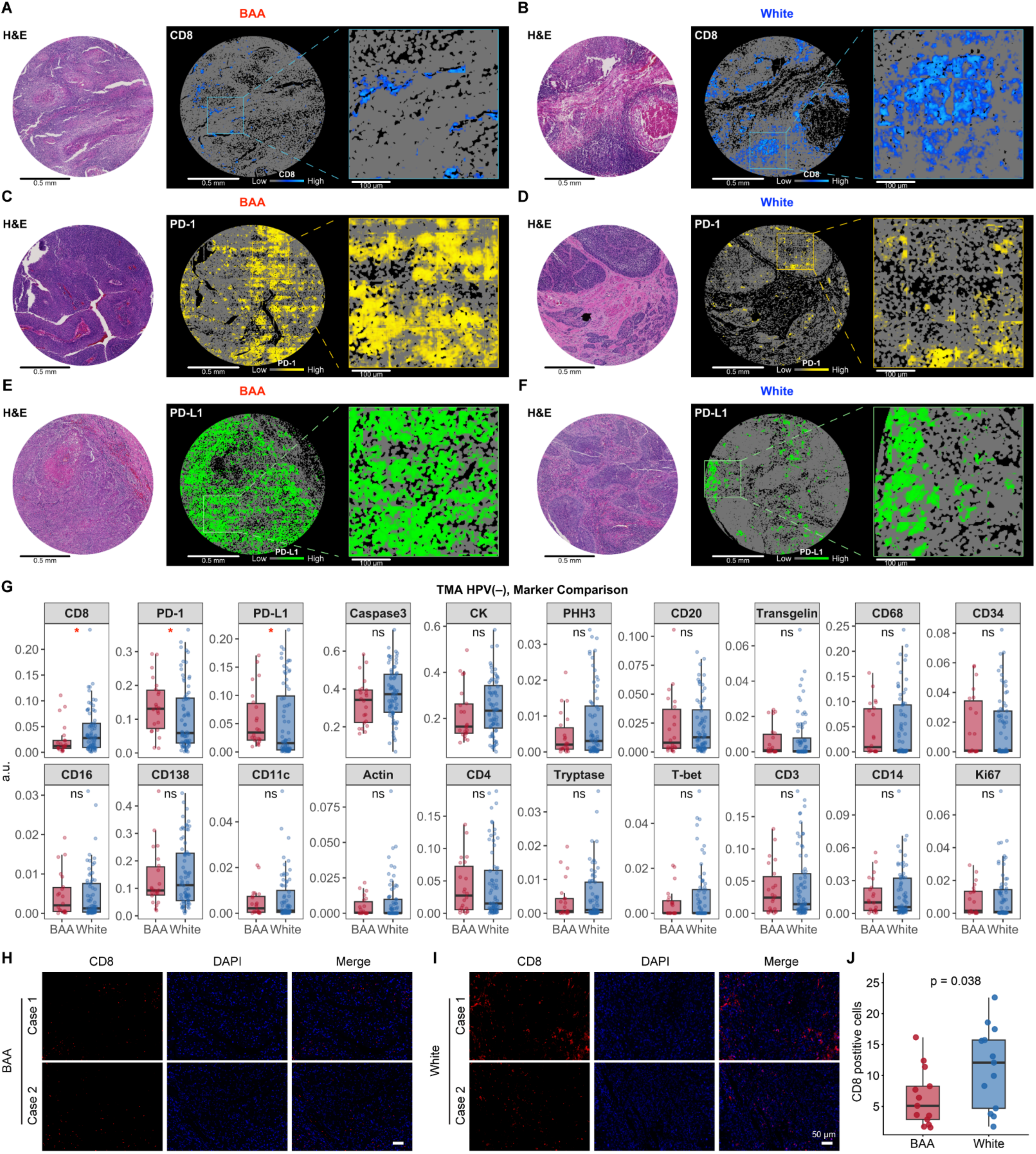
Validation of race-dependent immune differences in HPV-negative HNSCC in the Emory TMA cohort. (**A-F**) Representative H&E images, whole-slide virtual staining heatmaps, and magnified regions for CD8 (**A, B**), PD-1 (**C, D**), and PD-L1 (**E, F**) in BAA and White HPV-negative cases. Inset squares indicate regions shown at higher magnification. (**G**) Whole-slide mean expression of 20 virtual immunofluorescence markers comparing BAA and White HPV-negative HNSCC specimens from the Emory TMA cohort. (**H, I**) Representative CD8 immunofluorescence images from two BAA (**H**) and two White (**I**) HPV-negative tumors. (**J**) Quantification of CD8-positive cells comparing BAA and White-derived tumors (n = 13 per group). Scale bars, 0.5 mm (TMA cores), 100 μm (magnified regions), and 50 μm (immunofluorescence). Box plots show median and interquartile range with individual samples overlaid. ns, not significant, *p < 0.05, **p < 0.01, and ***p < 0.001, by Wilcoxon rank-sum test.

### Race-dependent differences in immune composition within the tumor core

To further characterize the spatial organization of these differences, we performed tissue niche segmentation, classifying image regions into tumor core, CAF/stroma, and immune-enriched niches. Niche identity was defined by marker enrichment profiles: the tumor core was characterized by high CK with low stromal and immune signal; the CAF/stroma niche showed elevated actin and transgelin, consistent with fibroblast-associated features, together with low CK and immune marker expression; and the immune-enriched niche was enriched for classical immune markers, including CD3, CD4, CD8, CD11c, CD14, CD16, CD20, and CD68 (**Fig. 4A-B**). Comparison of niche composition revealed a higher tumor core fraction in BAA-derived tumors (**Fig. 4C**). Importantly, analysis of immune marker expression within the tumor core showed that race-dependent differences in CD8, PD-1, and PD-L1 persisted within the tumor compartment (**Fig. 4D**), indicating that these disparities are not solely explained by differences in overall tissue composition but also reflect distinct patterns of immune infiltration and checkpoint expression within tumor regions. The same niche segmentation applied to the independent TMA cohort recapitulated the three compartments with comparable marker enrichment profiles (**Fig. 4E-F**). Consistent with the TCGA results, BAA TMA cores showed a higher tumor core fraction (**Fig. 4G**), and race-dependent differences in CD8, PD-1, and PD-L1 persisted within the tumor core (**Fig. 4H**), further supporting that these disparities reflect differences in immune infiltration and checkpoint expression within tumor regions in addition to overall tissue composition.

**Figure 4.**
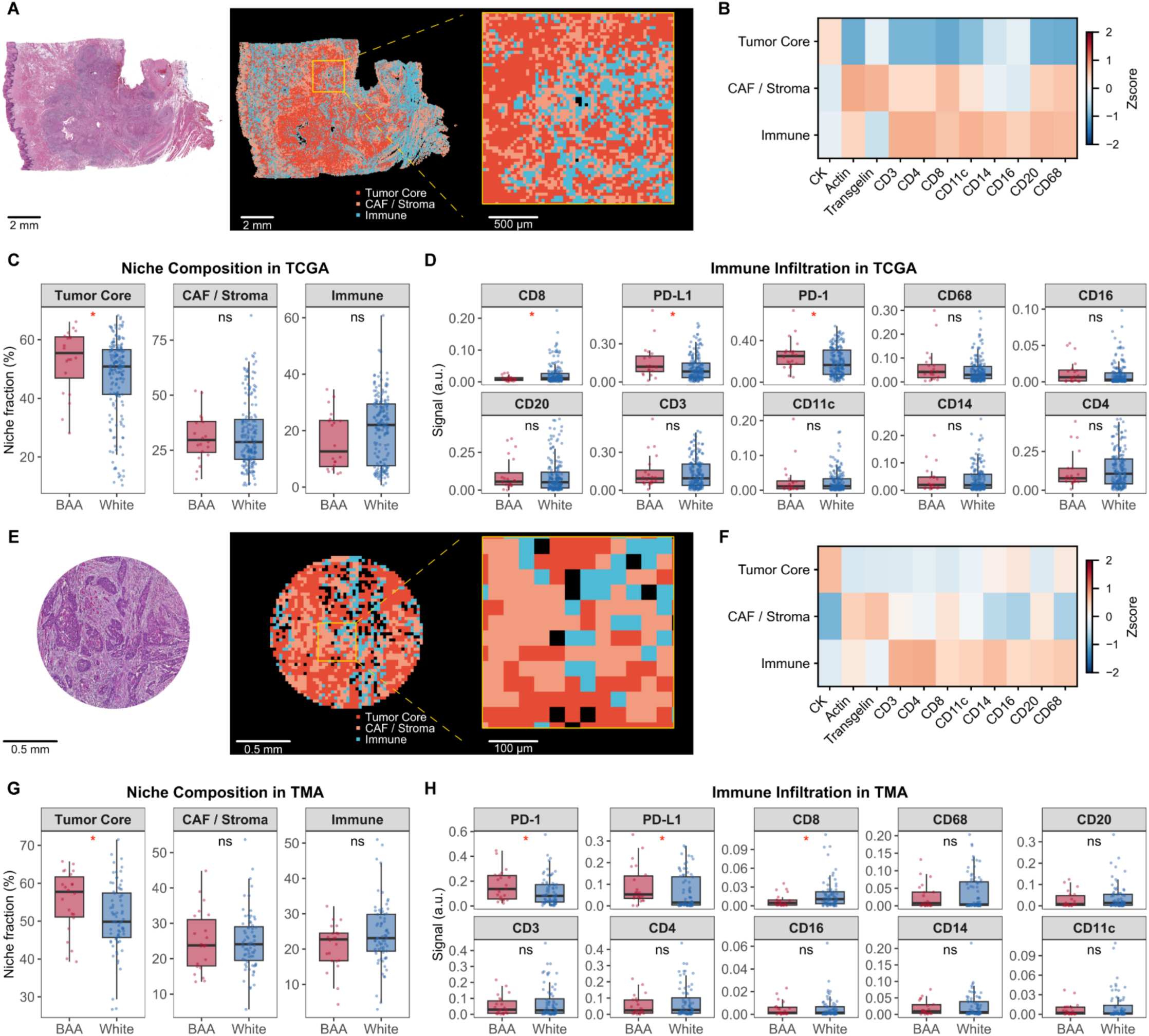
Tissue niche segmentation reveals race-dependent immune infiltration differences in HPV-negative HNSCC. (**A, B, E,** and **F**) Representative H&E image with tissue niche segmentation (tumor core, CAF/stroma, and immune compartments) in HNSCC specimens from the TCGA cohort (**A, B**) and the Emory TMA cohort (**E, F**). (**B**, **F**) Marker Z-score heatmaps across the three niches. (**C**, **G**) Niche composition comparing BAA and White-derived tumors. (**D, H**) Immune marker expression within the tumor core comparing BAA and White-derived tumors. Scale bars, 2 mm and 500 μm (**A**); 0.5 mm and 100 μm (**E**). Box plots show median and interquartile range with individual samples overlaid. ns, not significant, *p < 0.05, **p < 0.01, and ***p < 0.001 by Wilcoxon rank-sum test.

### MET emerges as a prominent race-associated gene elevated in BAA-derived HNSCC compared with White-derived HNSCC

Next, we applied Phoenix^19^, a deep learning model that infers spatial gene expression from H&E images, to further dissect the molecular basis of the immune disparities between BAA and White HNSCC (**Fig. 5A**). Tumor regions were defined by spots in the top quartile of a composite score from four epithelial markers (EPCAM, LY6D, COL17A1, KRT7), with distinct expression patterns across epithelial, CAF, and immune markers between high- and low-tumor regions (**Fig. 5B-C**). Predicted expression analysis within tumor regions across 379 HPV-negative TCGA H&E slides (BAA, n = 41; White, n = 338) revealed distinct race-associated molecular profiles (**Fig. 5D**). Compared with BAA-derived tumors, White-derived tumors showed a more immune “hot” profile, with T cell-recruiting chemokines CXCL9 and CXCL10 among the most highly upregulated genes (**Fig. 5D**), consistent with the immune differences observed in both the TCGA (**Fig. 2G**) and Emory TMA (**Fig. 4G**) cohorts. In contrast, MET, a receptor tyrosine kinase that promotes tumor growth, invasion, survival, and immune evasion, emerged as one of the genes most highly elevated in BAA-derived tumors compared with White-derived tumors (**Fig. 5D**). Notably, predicted MET expression was markedly higher in tumor tissues than in normal tissues in the TCGA HNSCC cohort (**Fig. S2**), demonstrating tumor-associated elevation of MET in HNSCC. Consistently, predicted tumoral MET expression in the Emory TMA HNSCC cohort was significantly higher in BAA-derived tumors than in White-derived tumors (BAA, n = 21; White, n = 73; p = 3.4 × 10^-3^; **Fig. 5E**). Spatial mapping further demonstrated higher MET expression in BAA-derived tumors in both the TCGA (**Fig. 5F-G**) and Emory TMA HNSCC cohorts (**Fig. 5H-I**). Moreover, immunofluorescence staining in HNSCC specimens obtained through CTP Shared Resource at Winship Cancer Institute of Emory University (BAA, n = 13; White, n = 13) confirmed significantly higher MET expression in BAA-derived tumors than in White-derived tumors (p = 0.041; **Fig. 5J-L**), further supporting MET as a racially enriched biomarker candidate in BAA HNSCC.

**Figure 5.**
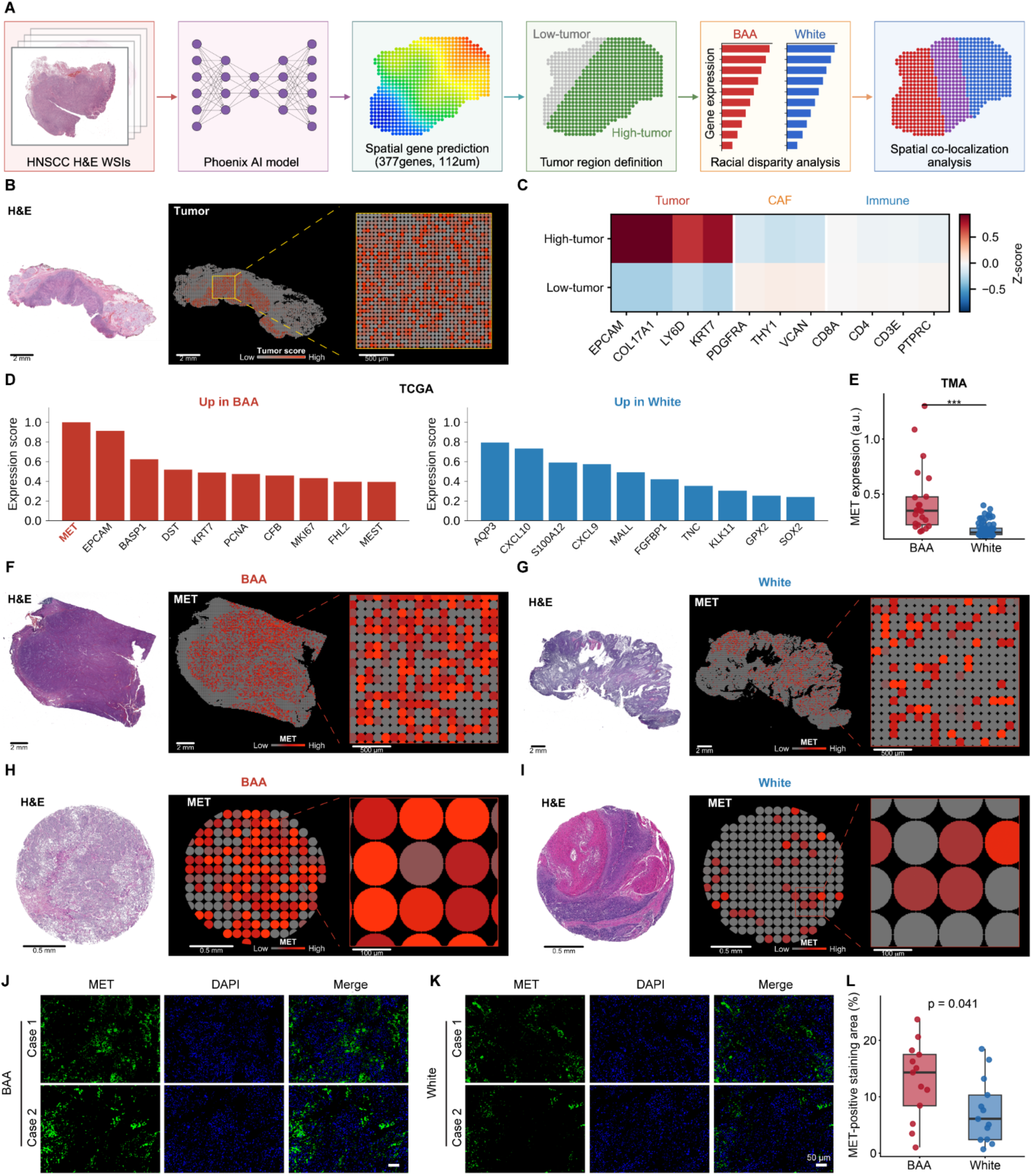
AI-based spatial transcriptomics identifies MET as a racially enriched biomarker in HPV-negative HNSCC. (**A**) Overview of the Phoenix analytical workflow. (**B**) Representative TCGA slide with H&E and Phoenix-predicted tumor score at spot-level resolution, with magnified region. (**C**) Marker expression Z-scores in high- and low-tumor regions across epithelial, CAF, and immune markers. (**D**) Top 10 differentially expressed genes upregulated in BAA (red) and White (blue) HPV-negative tumors from the TCGA HNSCC cohort. (**E**) Predicted MET expression in BAA versus White HPV-negative tumors from the Emory TMA HNSCC cohort. (**F-I**) Representative MET expression heatmaps with H&E, full overlay, and magnified regions in HNSCC specimens from the TCGA (**F**) and Emory TMA (**I**) cohorts. (**J, K**) Representative MET immunofluorescence images (MET, DAPI, and merged) from two BAA (**J**) and two White (**K**) tumors. (**L**) Quantification of the percentage of MET-positive staining area in BAA and White-derived tumors (n = 13 per group). Scale bars, 2 mm and 500 μm (TCGA); 0.5 mm and 100 μm (TMA); 50 μm (immunofluorescence). *p < 0.05, **p < 0.01, and ***p < 0.001, by Wilcoxon rank-sum test.

### MET-associated CD8 exclusion is more pronounced in BAA-derived HNSCC

We next examined whether MET expression was spatially associated with CD8⁺ T cell infiltration within the TME. Across 379 HPV-negative TCGA slides, MET-high regions showed significantly lower CD8 scores than MET-low regions (paired Wilcoxon test, p < 1 × 10^-3^; **Fig. 6A-B**), indicating spatial exclusion between MET expression and CD8 T cell infiltration. This pattern was consistent in the TMA cohort across 94 HPV-negative patients (paired Wilcoxon test, p < 1 × 10^-3^; **Fig. 6C-D**). When stratified by race, the inverse correlation between MET and CD8 was significant in both BAA and White patients but consistently stronger in BAA-derived tumors in both the TCGA cohort (BAA: r = -0.39, p = 0.012; White: r = -0.17, p = 0.002; **Fig. 6E**) and the Emory TMA cohort (BAA: r = -0.48, p = 0.023; White: r = -0.23, p = 0.050; **Fig. 6F**), suggesting that MET-associated immune exclusion is more pronounced in the BAA group. To experimentally validate this spatial relationship, we performed MET/CD8α co-IF staining in HNSCC specimens collected through CTP Shared Resource at the Winship Cancer Institute of Emory University (BAA, n = 13; White, n = 13). Consistent with the computational analysis, MET expression was significantly inversely correlated with CD8-positive cell infiltration across all patients (r = -0.59, P = 0.002; **Fig. 6G-H**), with a stronger association in BAA patients (r = -0.64, P = 0.019) than in White patients (r = -0.55, P = 0.049; **Fig. 6I**). These findings suggest that elevated MET expression may contribute to an immune-excluded TME in BAA HNSCC and provide a potential mechanistic link between race-associated MET enrichment and reduced CD8⁺ T cell infiltration.

**Figure 6.**
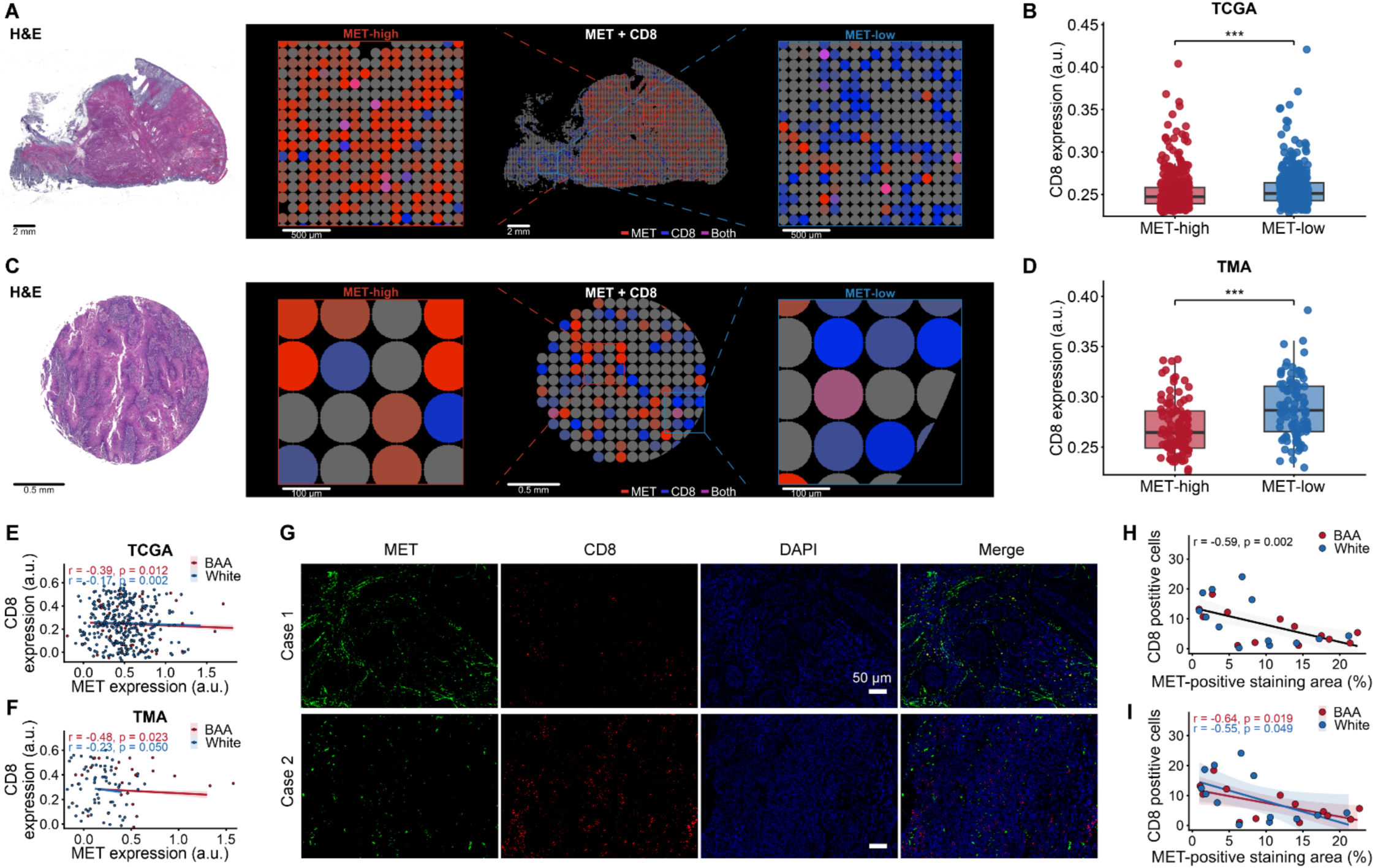
MET-CD8 spatial exclusion in HPV-negative HNSCC. (**A**) Representative TCGA whole-slide image with H&E and spatial expression overlays of MET and CD8. red = MET, blue = CD8, purple = co-expression. (**B**) CD8 expression in MET-high versus MET-low tumor regions across HNSCC specimens from the TCGA cohort. (**C**) Representative TMA core image with H&E and spatial expression overlays of MET and CD8. (**D**) CD8 expression in MET-high versus MET-low regions across HNSCC specimens from the Emory TMA cohort. (**E, F**) Correlation between MET and CD8 expression in HNSCC specimens stratified by race in the TCGA (**E**) and Emory TMA (**F**) cohorts. (**G**) Representative MET and CD8 co-immunofluorescence staining images from two different tumors. (**H, I**) Correlation between the percentage of MET-positive staining area and the number of CD8-positive cells detected by immunofluorescence across all cases (n = 26) (**H**) or stratified by race (n = 13 per group) (**I**). Scale bars, 2 mm and 500 μm (TCGA); 0.5 mm and 100 μm (TMA); 50 μm (immunofluorescence). ***p < 0.001, by paired Wilcoxon signed-rank tests in (**B, D**) and Pearson correlation coefficients in (**E**, **F**, **H**, **I**).

### Elevated MET expression may contribute to racial disparities in therapeutic response in HNSCC

To evaluate the clinical relevance of MET for treatment, we applied Phoenix to infer spatial gene expression from pretreatment H&E whole-slide images of HNSCC patients enrolled in a phase 2 trial of combined pembrolizumab (a PD-1 inhibitor) and cabozantinib (a MET-targeting multikinase inhibitor)^1,21^. Of the 17 patients evaluated, predicted MET expression in tumor regions was significantly higher in BAA than in White patients (BAA, n = 4; White, n = 13; p = 0.045; **Fig. 7A, B, G**), consistent with the racial disparity observed in the TCGA and Emory TMA cohorts. We next compared MET expression in tumor regions between patients with partial response (PR, n = 9) and stable disease (SD, n = 8) and found that PR patients had significantly higher predicted MET expression than SD patients (p = 0.027; **Fig. 7C, D, H**). In contrast, predicted PD-L1/CD274 expression did not differ significantly between PR and SD patient group (**Fig. S3**).

**Figure 7.**
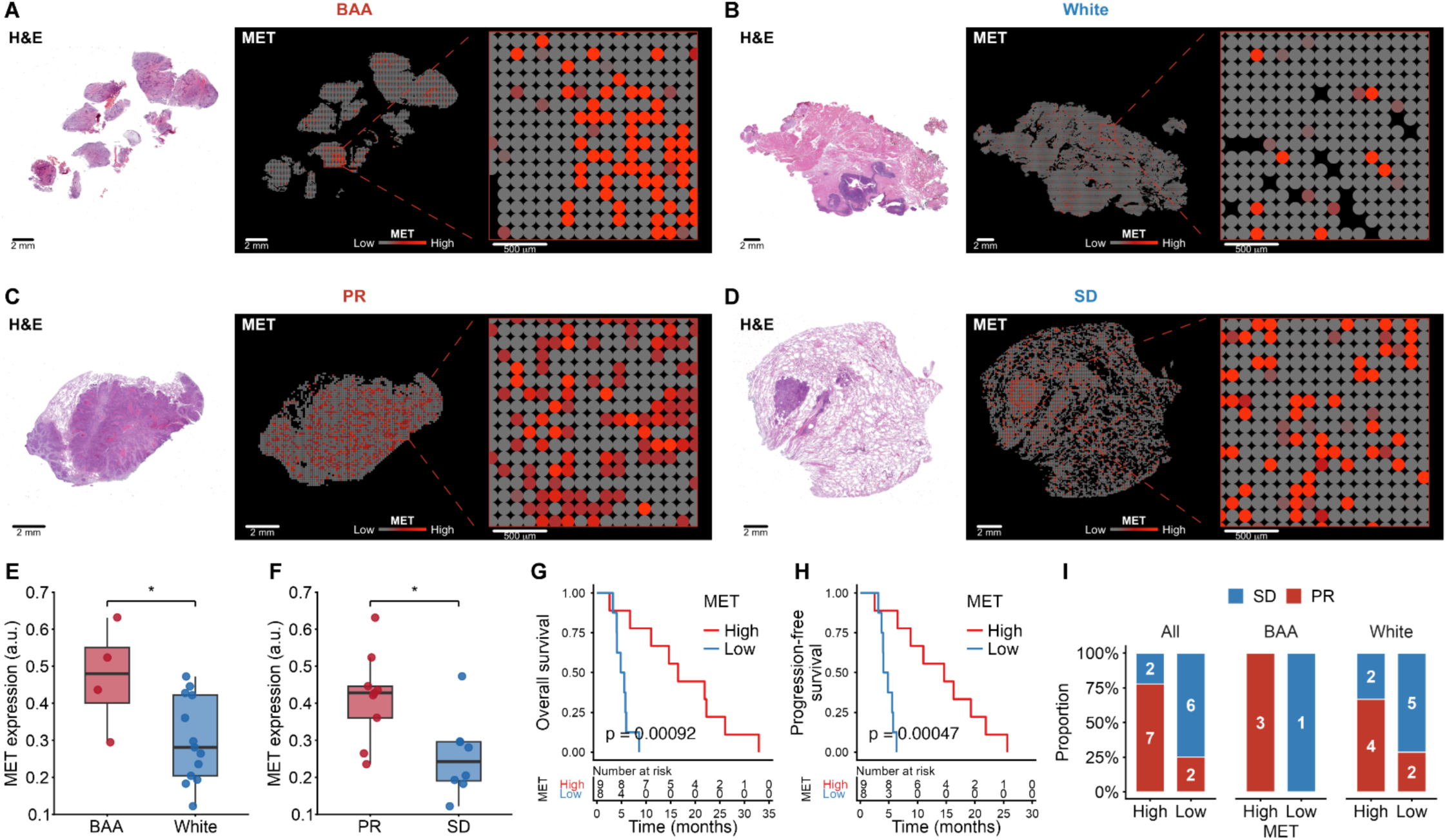
Predicted spatial MET expression and its correlation with treatment response in a phase 2 HNSCC trial cohort treated with pembrolizumab plus cabozantinib. (**A, B**) Representative HPV-negative BAA and White cases showing H&E whole-slide image, MET spatial heatmap, and magnified tumor region. (**C, D**) Representative partial response (PR) and stable disease (SD) cases showing MET spatial heatmap and magnified tumor region. (**E**) MET expression in tumor regions comparing BAA (n = 4) versus White (n = 13) patients. (**F**) MET expression in tumor regions comparing PR (n = 9) versus SD (n = 8) patients. (**G**) Correlation between MET expression and overall survival in HNSCC patients treated with combined pembrolizumab and cabozantinib. (**H**) Correlation between MET expression and progression-free survival in HNSCC patients treated with combined pembrolizumab and cabozantinib. (I) Proportions of HNSCC patients with PR or SD among MET-high and MET-low groups in the overall cohort, BAA patients, and White patients. Numbers within bars indicate patient counts. Scale bars, 2 mm and 500 μm. *p < 0.05 by Wilcoxon rank-sum test.

Moreover, higher MET expression was significantly associated with longer overall survival (median 16.5 vs. 5.2 months; log-rank p = 0.00092) and progression-free survival (median 14.6 vs. 4.4 months; log-rank p = 0.00047) in patients treated with combined pembrolizumab and cabozantinib (**Fig. 7G-H**). Notably, when patients were stratified according to the median MET expression level, 78% of MET-high patients achieved a partial response (PR), compared with only 25% of MET-low patients (**Fig. 7I**). When further stratified by race, all BAA patients in the MET-high group achieved a PR, compared with none of the BAA patients in the MET-low group (100% vs. 0%) (**Fig. 7I**). Among White patients, 67% of those in the MET-high group achieved PR compared with 29% in the MET-low group (**Fig. 7I**). These findings further support MET as a potential predictive biomarker of clinical benefit from pembrolizumab plus cabozantinib therapy, particularly among BAA patients, and may provide insight into racial disparities in therapeutic response in HNSCC.

## DISCUSSION

The findings of the present study nominate MET-associated immune exclusion as a spatially resolved candidate mechanism underlying racial differences in the HNSCC tumor immune microenvironment. The CD8-low, checkpoint-high phenotype observed in BAA-derived tumors defines an immunologically unfavorable profile resembling the immune-excluded phenotype associated with limited responsiveness to immune checkpoint blockade across multiple cancer types^22,23^. Critically, this pattern persists within the tumor core rather than simply reflecting intratumoral heterogeneity or differences in tissue composition, suggesting that conventional tissue-level immune scoring may underestimate the extent of immune exclusion in BAA-derived tumors. Interestingly, MET provides a molecular candidate connecting this exclusionary phenotype to a potentially targetable pathway. Although MET has well-characterized roles in immune evasion, including dendritic cell suppression, resistance to EGFR-based therapy, PD-L1 upregulation, and myeloid reprogramming^24–28^, these functions have not previously been linked to race-dependent spatial organization of the immune microenvironment. The spatial inverse correlation between MET and CD8 within individual tumors, and its amplification in BAA-derived tumors, further raises the possibility that MET contributes to the organization of an immune-excluded tumor architecture rather than simply co-occurring with an immune-cold state.

Our findings also provide a rationale for therapeutic strategies that simultaneously target tumor-intrinsic immune exclusion and adaptive immune checkpoint signaling^23,29,30^. In this context, combining MET inhibition with PD-1 blockade may be particularly advantageous. While PD-1 inhibition primarily releases inhibitory signals that restrain existing antitumor T cell activity and may be insufficient in tumors characterized by limited T cell infiltration^22,29^, MET inhibition has the potential to remodel the TME by reducing immunosuppressive myeloid populations, attenuating pro-angiogenic signaling, and promoting CD8⁺ T cell recruitment^31–35^. Importantly, this therapeutic concept has already been explored clinically by our group in recurrent/metastatic HNSCC^1,21^. Our phase 2 trial of pembrolizumab plus cabozantinib reported an objective response rate of 52% among 33 evaluable patients, with the median progression-free survival and overall survival 14.6 and 22.3 months, respectively^1^. Longer-term follow-up reported a median overall survival of 27.7 months and a 2-year overall survival of 54.7%^21^. We have previously reported that baseline phosphorylated MET levels were associated with treatment response, supporting a potential role for MET signaling in determining benefit from pembrolizumab plus cabozantinib therapy^21^. Here, we extend these observations by showing that higher predicted spatial MET expression is strongly associated with improved treatment response. Notably, this association was more pronounced among BAA patients, suggesting that MET may be particularly informative in this population and providing a potential link between the race-associated spatial biology observed in our cohorts and differential therapeutic benefit. However, validation in larger, racially balanced cohorts and randomized clinical studies will be necessary to establish the predictive value of MET and determine whether MET identifies patients who derive particular benefit from the addition of cabozantinib to PD-1 blockade. As other MET targeted approaches are in use in HNSCC namely to overcome EGFR resistance mechanisms, further exploration of these associations may be relevant in these clinical settings^36,37^.

The H&E-based computational approach demonstrated here could be extended to other cancer types where racial outcome disparities and MET dysregulation have been documented, including triple-negative breast cancer (TNBC), prostate cancer, and colorectal cancer^38–43^. A recent spatial multi-omics study of TNBC identified racially distinct multicellular niches and immunosuppressive mechanisms, with BAA-derived tumors exhibiting an immune-cold, endothelial-macrophage-mesenchymal niche architecture^40^. Whether MET contributes to similar exclusionary phenotypes in these contexts remains an open question that deserves investigation. As archival H&E slides are widely available across cancer types and institutions, this framework enables investigation of race-associated microenvironmental features at a scale impractical with conventional spatial technologies such as multiplex immunofluorescence, imaging mass cytometry, or sequencing-based spatial transcriptomics^44–46^. This scalability is a critical advantage for health disparities research, where the most important cohorts often have limited remaining tissue and where the underrepresentation of minority patients in existing spatial profiling datasets has left fundamental questions about race-associated tumor biology unanswered^40,47,48^. Beyond scalability, a key strength of GigaTIME and Phoenix is their ability to infer complementary cellular and molecular features directly from H&E morphology without requiring additional assays. Their pan-cancer training supports broad applicability. Notably, Phoenix accurately predicted compartment-specific gene expression, including MET, in an HNSCC lymph-node metastasis that was excluded from model training^19^, supporting the feasibility of applying this approach to the present HNSCC cohorts. Nevertheless, AI-derived spatial predictions should be interpreted as computationally inferred measurements rather than direct experimental measurements, and prediction accuracy may vary across genes, cell types, tissue compartments, and histologic contexts. Although the observed MET-associated spatial patterns were supported by independent MET immunofluorescence validation, broader orthogonal validation across additional markers and cohorts will be important to establish the robustness and generalizability of these predictions. Future fine-tuning using larger, demographically diverse, HNSCC-specific spatial-transcriptomic datasets may further improve cancer-specific performance and reduce potential biases. In addition, standardized stain and scanner harmonization, together with cross-institutional calibration, will be important for improving reproducibility and facilitating translation of these AI-based spatial profiling approaches into multicenter studies.

Our findings are encouraging, but several important questions remain to be addressed. In particular, the relatively modest representation of BAA patients in both cohorts (the TCGA HPV-negative cohort, n = 41; the Emory TMA HPV-negative cohort, n = 22) highlights the need for larger and more diverse patient databases. Future prospective studies with intentional and enhanced racial representation will be essential to validate these observations, better define the molecular and immune features underlying racial disparities in HNSCC, and determine their broader clinical relevance^49^. The clinical cohort was small (n = 17) and single-arm, so the observed response and survival associations should be considered exploratory and require validation in larger independent studies. By restricting race comparisons to HPV-negative disease, we reduced potential confounding by HPV status; future multivariable analyses incorporating age, smoking exposure and treatment history will help determine the independence of the observed race-associated differences. In addition, the spatial transcriptomic inference covers 377 genes rather than the full transcriptome, and additional candidates relevant to race-dependent immune exclusion may exist outside this panel; unbiased spatial transcriptomic profiling in future studies could reveal additional targets beyond MET. All expression values are computationally predicted, and although cross-cohort concordance and experimental validation of MET provide orthogonal support for the central findings, systematic comparison with ground-truth multiplex spatial data will be important for more comprehensive validation^50^. The current study is cross-sectional and establishing MET association with immune exclusion will require functional investigation in preclinical models. Moreover, race is a social construct encompassing genetic ancestry, environmental exposures, and structural determinants. These findings characterize tumor-level biological differences but do not diminish the central role of systemic factors in driving HNSCC disparities^51–53^.

In summary, our study provides spatial and clinical evidence that MET is closely linked to immune exclusion and therapeutic response in HNSCC and may represent a molecular feature contributing to racial disparities in disease biology and treatment benefit. The convergence of elevated MET expression, diminished CD8⁺ T cell infiltration, and enhanced response to pembrolizumab plus cabozantinib among MET-high tumors, particularly in BAA patients, suggests that MET-associated biology may define a therapeutically actionable tumor state. These findings position MET as a promising candidate biomarker for patient stratification and provide a rationale for investigating MET-directed strategies to overcome immune exclusion and potentially narrow racial disparities in HNSCC outcomes.

## MATERIALS AND METHODS

### The TCGA HNSCC cohort and data acquisition

H&E-stained whole-slide images from the TCGA HNSCC cohort were downloaded from the Genomic Data Commons (GDC) Data Portal (https://portal.gdc.cancer.gov)^20^. A total of 472 FFPE slides were retrieved after excluding frozen sections. After removing duplicate slides, 450 unique cases were retained. An additional 42 normal tissue slides available from the TCGA-HNSCC cohort were also downloaded for comparison. Clinical metadata, including race, TNM stage, sex, age, and smoking history, were obtained from the TCGA clinical metadata. HPV status was assigned based on the classification by Nulton et al^54^. Cases were stratified as HPV-positive (n = 46) or HPV-negative (n = 404), with racial groups (BAA and White) defined by the TCGA clinical metadata.

### The Emory TMA HNSCC cohort and TMA construction

All clinical specimens were collected with written informed consent under an Emory IRB-approved protocol (IRB00003208). HNSCC cases from 2015-2022 were selected from the Emory University Department of Pathology clinical archive and included pretreatment biopsy or surgical specimens. Diagnoses were independently confirmed by at least two pathologists. Clinical data, including race, demographic information, TNM stage, lymph node status, smoking history, HPV status, treatment history, and survival outcomes, were maintained in Emory WinDATA in compliance with HIPAA regulations. Diagnostic H&E slides were reviewed to identify representative tumor and normal regions for TMA construction. Triplicate 1.5-mm cores from each tumor, along with representative normal tissue cores, were transferred into recipient TMA blocks using a semi-automated tissue microarrayer. TMA sections (5 μm) were stained with H&E and digitally scanned using the Olympus Nanozoomer whole-slide scanner (Olympus, Center Valley, PA).

### A phase 2 HNSCC trial cohort treated with combined pembrolizumab and cabozantinib

Patients with recurrent or metastatic HNSCC were enrolled in a phase 2, single-arm clinical trial evaluating pembrolizumab plus cabozantinib (ClinicalTrials.gov, NCT03468218, registered March 16, 2018) at the Winship Cancer Institute of Emory University between March 2019 and June 2022. Eligible patients had measurable disease according to RECIST v1.1 and no contraindications to either treatment. Among the 36 patients enrolled in the trial, 33 were evaluable for treatment response. Of these, 17 patients, including 4 BAA and 13 White patients, had pretreatment H&E-stained whole-slide images with corresponding clinical annotations available for the present spatial analysis. Best overall response was assessed according to RECIST v1.1. Clinical data, including race, treatment response, HPV status, and PD-L1 combined positive score (CPS), were obtained from the clinical trial database, which has been previously described in detail.^1^

### GigaTIME virtual mIF

Whole-slide H&E-stained images from 450 TCGA-HNSCC FFPE tumor sections with matched clinical metadata were obtained from the GDC; frozen sections were excluded to ensure optimal virtual staining performance. Virtual mIF was performed using GigaTIME, an artificial intelligence framework that employs a deep learning architecture to predict spatial protein expression maps directly from H&E-stained whole-slide images. Each whole-slide image was tiled into 256 × 256 pixel patches, tiles with less than 20% tissue content were excluded, and the remaining tiles were processed with GigaTIME to generate virtual immunofluorescence images for 20 protein markers (Actin, CD11c, CD138, CD14, CD16, CD20, CD3, CD34, CD4, CD68, CD8, CK, Caspase-3, Ki67, PD-1, PD-L1, PHH3, T-bet, Transgelin, and Tryptase); predicted tiles were reassembled into whole-slide marker maps based on tile coordinates. A tissue mask was derived from the virtual DAPI channel by intensity thresholding, and whole-slide mean intensity was calculated in arbitrary units (a.u.) for each marker across all tissue-masked pixels. For spatial niche analysis, tissue regions were classified into tumor core, CAF/stroma, and immune-enriched niches based on marker enrichment profiles, and immune infiltration within the tumor core was quantified as the mean immune marker intensity within tumor core regions. The same pipeline was applied to nine tissue microarrays from the Emory Winship cohort; individual cores were isolated using circular masks, and per-patient values were obtained by averaging across cores. Differences in marker intensity and niche composition were assessed using the two-sided Wilcoxon rank-sum test (p < 0.05). Representative spatial maps were visualized with continuous color scales on a black background, and all figures were generated using R (v4.x) with the png package.

### GigaTIME-based immune risk score development

To construct an immune-based risk score from GigaTIME data, we applied LASSO Cox proportional hazards regression to the mean fluorescence intensity values of all 21 biomarker channels. GigaTIME virtual multiplex immunofluorescence data were available for 450 TCGA-HNSC patients; after excluding 3 patients lacking overall survival information, 447 patients were included in the analysis. Channel intensities were standardized to zero mean and unit variance prior to model fitting. The optimal regularization parameter (α) was selected via 5-fold cross-validation by maximizing Harrell’s concordance index (C-index), yielding a cross-validated C-index of 0.549 (SD = 0.022) at α = 0.0013. The final model retained all 21 markers with non-zero coefficients. For each patient, the risk score was computed as the linear combination of standardized marker intensities weighted by the corresponding LASSO Cox coefficients. Patients were stratified into high- and low-risk groups using the median risk score as the cutoff. The model was trained on the entire cohort and subsequently evaluated in HPV-negative (n = 401) and HPV-positive (n = 46) subgroups. Overall survival differences between risk groups were assessed by the log-rank test, and hazard ratios (HR) with 95% confidence intervals were estimated using Cox regression. Risk score distributions between racial groups (BAA vs. White) were compared using the Wilcoxon rank-sum test.

### Phoenix spatial gene expression prediction

Spatial gene expression was inferred from H&E images using Phoenix, a deep learning model that predicts transcript-level expression for 377 genes at 112 µm spot-level resolution, matching the spatial resolution of the 10x Visium platform. For each spot, a tissue patch centered at the spot location was extracted from the H&E image and passed through the model to generate predicted expression values for all 377 genes. For the TCGA cohort, 379 HPV-negative FFPE whole-slide images were processed. For the Emory TMA cohort, nine TMA whole-slide images were processed; individual spots were assigned to TMA cores based on Euclidean distance to core centers derived from the TMA grid layout, and per-patient values were obtained by averaging predicted expression across all spots within matched cores. All predicted expression values are reported in arbitrary units as output by the model.

### Tumor region definition and racial differential expression

Tumor regions were defined at the spot level using a composite score computed as the mean predicted expression of four epithelial markers (EPCAM, LY6D, COL17A1, KRT7). Spots in the top quartile of this composite score were classified as tumor spot. Differences in predicted expression between BAA and White patients were assessed by computing the mean predicted expression of each gene across tumor spots per patient, followed by a two-sided Wilcoxon rank-sum test. Genes were ranked by the absolute difference in group means (BAA versus White), and the top 10 genes with higher expression in each group were reported. For visualization, expression differences were normalized to the maximum absolute difference across all genes.

### MET-CD8 spatial exclusion analysis

To assess the spatial relationship between MET expression and CD8 T cell infiltration, tumor spots within each slide or TMA patient were ranked by predicted MET expression. Spots in the top 25% were classified as MET-high and the bottom 25% as MET-low. A CD8 score was computed for each spot as the mean predicted expression of CD8A and CD3E. The mean CD8 score was compared between MET-high and MET-low regions within each patient using a paired Wilcoxon signed-rank test. This analysis was performed independently in 373 TCGA slides and 94 HPV-negative TMA patients. Spatial overlay maps were generated by combining MET (red channel) and CD8 (blue channel) predicted signals, with co-expression rendered in purple.

### Visualization

Spatial expression maps were rendered at 8× downsampled resolution on a black background, with predicted expression values mapped to continuous red (MET) or blue (CD8) color scales. For MET-CD8 overlay maps, red and blue channels were combined, with co-expression regions rendered in purple. Zoomed regions (2 mm × 2 mm for TCGA, 0.4 mm × 0.4 mm for TMA) were selected to illustrate areas of highest MET or CD8 signal. TMA core boundaries were clipped using circular masks derived from core center coordinates. Boxplots show median and interquartile range with individual data points overlaid. Bar plots of differential expression display normalized expression differences scaled to the maximum value. All figures were generated using R (v4.x) with ggplot2 and the png package.

### Immunofluorescence (IF)

IF was carried out following a previously established protocol^55^. In brief, tumor tissue was fixed with 4% paraformaldehyde for 15 min and permeabilized with 0.1% Triton X-100 for 10 min. After blocking for 1 h, tumor sections were incubated with CD8α rabbit monoclonal antibody (Cell signaling technology, Cat #85336) and/or MET mouse monoclonal antibody (Cell signaling technology, Cat #8741) at 4 °C overnight, followed by incubation with goat anti-rabbit Alexa Fluor 594-conjugated secondary antibody (TheromFisher, Cat #A-11037) and/or goat anti-mouse Alexa Fluor 488-conjugated secondary antibody (TheromFisher, Cat #A-11029) in the dark for 1 h. Slides were mounted with Vectashield mounting medium containing DAPI (Vector Laboratories, Burlingame, CA) and visualized using an inverted fluorescence microscope (BZ-X710 All-in-one, Keyence). Quantitation of CD8-positive cells in tissue section was counted in six randomly chosen fields, and the average number of positive cells per reported field was calculated based on the results provided by two investigators. Quantification of MET expression was performed by analyzing the positive percentage of staining area in six randomly chosen fields per tissue section using the Image J software (NIH).

### Survival analysis

Associations between tumor MET expression and clinical outcomes were assessed by stratifying patients into MET-high and MET-low groups using the median expression value as the cutoff. Progression-free survival and overall survival were estimated using the Kaplan-Meier method, and differences between groups were evaluated using the log-rank test. Overall and progression-free survival were estimated by the Kaplan-Meier method; patients without an event at the data cutoff were censored at the date of last follow-up. Survival analyses were performed in R using the survival and survminer packages.

### Statistical analysis

All statistical comparisons between two groups were performed using the two-sided Wilcoxon rank-sum test for unpaired data or the paired Wilcoxon signed-rank test for within-patient comparisons (e.g., MET-high versus MET-low regions). P values less than 0.05 were considered statistically significant. No correction for multiple comparisons was applied to individual marker or gene-level tests. All analyses were performed using Python (v3.10) with NumPy and SciPy, and R (v4.x) with ggplot2. Sample sizes are reported in the corresponding figure legends and results text.

## Data Availability

All data produced in the present study are available upon reasonable request to the authors

## RESOURCE AVAILABILITY

### Lead contact

Further information and requests for resources and reagents should be directed to and will be fulfilled by the lead contact, Yong Teng

### Materials availability

No new materials were generated in this study; all materials are commercially available.

### Data and code availability

Data and code generated from this study will be made available upon request from the lead contact upon reasonable request and with a data use agreement.

## ACKNOWLEDGEMENTS

We would like to acknowledge the Winship Cancer Institute/Emory University cores that support this research, including Cancer Tissue and Pathology Shared Resource and Biostatistics Shared Resource (NIH/NCI award number P30CA138292).

## AUTHOR CONTRIBUTIONS

Project design was done by YT. Experiments were conducted by XC and FC. Data analysis and interpretation were contributed by XC, FC, NFS and YT. Patient case samples and details were provided by AM, YL, NCS, CHC, CES, DMS and NFS. Project supervision and funding was from YT. Manuscript writing was completed by XC and YT. Manuscript editing was completed by all authors.

